# Protection afforded by post-infection SARS-CoV-2 vaccine doses: a cohort study in Shanghai

**DOI:** 10.1101/2024.01.09.24301069

**Authors:** Bo Zheng, Bronner Gonçalves, Pengfei Deng, Weibing Wang, Jie Tian, Xueyao Liang, Ye Yao, Caoyi Xue

**Affiliations:** Department of Epidemiology, School of Public Health, Fudan University, Shanghai 200032, China; Department of Comparative Biomedical Sciences, School of Veterinary Medicine, University of Surrey, Guildford, UK; Shanghai Pudong New Area Center for Disease Control and Prevention, Shanghai, 200120, China; Key Laboratory of Public Health Safety of Ministry of Education, Fudan University, Shanghai 200032, China; School of Public Health, Shanghai Institute of Infectious Disease and Biosecurity, Fudan University, Shanghai 200032, China

**Keywords:** COVID-19, vaccine effectiveness, hybrid immunity, reinfection, Omicron variant

## Abstract

**Background:** In many settings, a large fraction of the population has both been vaccinated against and infected by SARS-CoV-2. Hence, quantifying the protection provided by post-infection vaccination has become critical for policy. We aimed to estimate the protective effect against SARS-CoV-2 reinfection of an additional vaccine dose after an initial Omicron variant infection.

**Methods:** We report a retrospective, population-based cohort study performed in Shanghai, China, using electronic databases with information on SARS-CoV-2 infections and vaccination history. We compared reinfection incidence by post-infection vaccination status in individuals initially infected during the April-May 2022 Omicron variant surge in Shanghai and who had been vaccinated before that period. Cox models were fit to estimate adjusted hazard ratios (aHR).

**Results:** 275,896 individuals were diagnosed with RT-PCR-confirmed SARS-CoV-2 infection in April-May 2022; 199,312/275,896 were included in analyses on the effect of a post-infection vaccine dose. Post-infection vaccination provided protection against reinfection (aHR 0.82; 95% CI 0.79-0.85). For patients who had received one, two or three vaccine doses before their first infection, hazard ratios for the post-infection vaccination effect were 0.84 (0.76-0.93), 0.87 (0.83-0.90) and 0.96 (0.74-1.23), respectively. Post-infection vaccination within 30 and 90 days before the second Omicron wave provided different degrees of protection (in aHR): 0.51 (0.44-0.58), and 0.67 (0.61-0.74), respectively. Moreover, for all vaccine types, but to different extents, a post-infection dose given to individuals who were fully vaccinated before first infection was protective.

**Conclusions:** In previously vaccinated and infected individuals, an additional vaccine dose provided protection against Omicron variant reinfection. These observations will inform future policy decisions on COVID-19 vaccination in China and other countries.

## 1 Introduction

Four years after the first reports of severe acute respiratory syndrome coronavirus 2 (SARSLJCoVLJ2) infection, the Coronavirus disease (COVID-19) pandemic continues to be a global concern, especially due to the risk of emergence of new variants[1, 2]. In most countries, the variant that is currently epidemiologically dominant is the Omicron[3, 4], which, due to its increased transmissibility and high number of mutations, led to significant increases in the number of infections in 2022[5]. Omicron variant infections were first observed in China in December 2021[3], and in Shanghai, the spread of the Omicron BA.2 sublineage led to a substantial increase in COVID-19 incidence between February 26 and June 30, 2022[6].

In December 2022[7], an important change in the COVID-19 policy in China, namely the end of most social distancing measures and of mass screening activities, was associated with a second surge in SARS-CoV-2 infections in Shanghai. The current circulation of the virus in the Shanghainese population and reports of vaccine fatigue mean that it is important to estimate the protective effect of vaccination against reinfection in this population. In this study, we aimed to quantify the effect of vaccine doses given after a first infection on the risk of subsequent infection. For that, we used data collected during the first Omicron variant wave, when hundreds of thousands of individuals tested real-time polymerase chain reaction (RT-PCR)-positive for SARS-CoV-2 infection[8] in Shanghai, of which 275,896 individuals in Pudong District. The fact that the population in Shanghai was mostly SARS-CoV-2 infection naïve before the spread of the Omicron variant provides a unique opportunity to estimate the real-world benefit of post-infection vaccine doses in a population that was first exposed to infection during a relatively short and well-defined time window. We further investigated whether the number of pre-infection vaccination doses modified the protective effect of the post-infection dose against Omicron BA.5 sublineage. To avoid ambiguity in the text, in the following sections, we often refer to vaccine doses given after the initial infection as “post-infection vaccination” or “post-infection vaccine dose”.

## 2 Results

### 2.1 Demographic characteristics of the study population

Of the 275,896 individuals with RT-PCR-confirmed SARS-CoV-2 infection during the first Omicron variant wave (from April 1 to May 31, 2022) in Pudong, Shanghai, 1,227 individuals died from reasons unrelated to COVID-19 between the first infection and November 2022 and were excluded from our analysis. Most first infections (243,906, 88.8%) occurred in April; for 306 (0.1%) individuals, information on the date of first infection was not available. In April 2022, more than half of the study population (68.6%) had completed full vaccination and one third (34.4%) had received booster vaccination.

To assess the effect of post-infection vaccination, the analytic sample consisted of 199,312 individuals (**Figure 1**). 85,804 were women (43.1%); 836 (0.4%) had sex information missing. 38.1% of the study participants were aged 20 to 39 years and only 0.9% were aged 0 to 6 years (see **Table 1** and **Table S1** for additional information).

**Figure 1.**
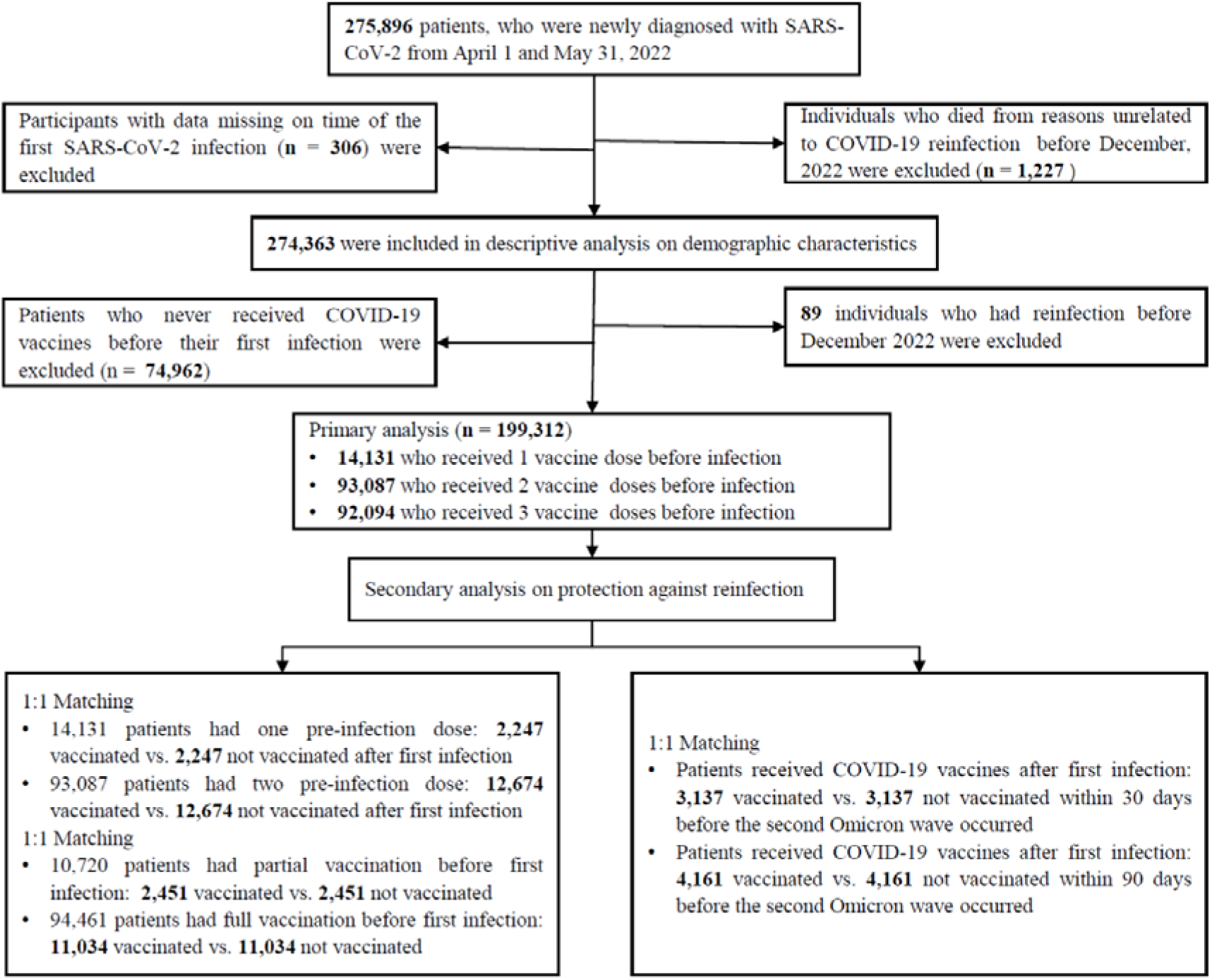
Flow chart describing the selection of participants for the analysis. The number of individuals in this figure is not the same as some of the numbers in Table 1 because of missing data in key variables. Note that in the bottom part of the chart, related to secondary analyses, the boxes represent overlapping sets of study participants; in other words, some individuals included in the secondary analyses that correspond to the left box were also included in analyses corresponding to the box on the right.

**Table 1.** Characteristics of the study population and reinfection rate by post-infection vaccination status. Here, reinfection rate refers to the percentage of the relevant study subpopulation with evidence of reinfection between December 1, 2022 and January 3, 2023. Note that for the variables on region, occupation, and clinical severity, data are missing for large fractions of the study population. Note also that information was only available on sex at birth, but not on gender.

| Characteristics |  | All |  | No post-infection vaccination |  | Post-infection vaccination |  |
| --- | --- | --- | --- | --- | --- | --- | --- |
|  |  | N (%) | Reinfection rate, %<br>(95% CI). | N (%) | Reinfection rate, %<br>(95% CI). | N (%) | Reinfection rate, %<br>(95% CI). |
| **Overall** |  | 199,312 | 24.4 (24.2, 24.6) | 183,165 | 24.7 (24.5, 24.9) | 16,147 | 21.5 (20.8, 22.2) |
| **Sex** | Male | 112,672 (56.5) | 26.1 (25.8, 26.4) | 104,002 (56.8) | 26.4 (26.1, 26.7) | 8,670 (53.7) | 22.3 (21.3, 23.3) |
|  | Female | 85,804 (43.1) | 22.4 (22.1, 22.7) | 78,403 (42.8) | 22.9 (22.6, 23.2) | 7,401 (45.8) | 17.4 (16.4, 18.3) |
| **Age, years** | 0-6 | 1,736 (0.9) | 7.0 (5.8, 8.3) | 1,569 (0.9) | 6.6 (5.4, 8.0) | 167 (1.0) | 10.2 (6.2, 15.9) |
|  | 7-19 | 10,762 (5.4) | 13.0 (12.3, 13.7) | 10,347 (5.6) | 12.9 (12.3, 13.6) | 415 (2.6) | 14.7 (11.4, 18.8) |
|  | 20-39 | 75,955 (38.1) | 22.4 (22.1, 22.8) | 71,005 (38.8) | 22.7 (22.3, 23.0) | 4,950 (30.7) | 19.1 (17.9, 20.3) |
|  | 40-59 | 74,680 (37.5) | 29.4 (29.0, 29.8) | 70,569 (38.5) | 29.6 (29.2, 30.0) | 4,111 (25.5) | 25.8 (24.2, 27.4) |
|  | 60+ | 35,903 (18.0) | 22.6 (22.1, 23.1) | 29,446 (16.1) | 23.7 (23.1, 24.2) | 6,457 (40.0) | 17.6 (16.6, 18.6) |
| **Regions** | Shanghai | 44,259 (22.2) | 18.9 (18.5, 19.3) | 41,250 (22.5) | 19.4 (19.0, 19.9) | 3,009 (18.6) | 11.9 (10.7, 13.1) |
|  | Other provinces | 44,959 (22.6) | 20.9 (20.5, 21.3) | 43,045 (23.5) | 21.0 (20.6, 21.5) | 1,914 (11.9) | 18.2 (16.4, 20.2) |
| **Occupations** | Preschoolers and students | 12,232 (6.1) | 12.1 (11.5, 12.7) | 11,677 (6.4) | 12.0 (11.4, 12.6) | 555 (3.4) | 13.2 (10.4, 16.4) |
|  | Employed | 29,537 (14.8) | 20.8 (20.2, 21.3) | 28,343 (15.5) | 20.8 (20.3, 21.4) | 1,194 (7.4) | 18.8 (16.4, 21.3) |
|  | Retired | 37,482 (18.8) | 22.5 (22.0, 23.0) | 30,955 (16.9) | 23.5 (23.0, 24.1) | 6,527 (40.4) | 17.5 (16.5, 18.5) |
|  | Working age not in labor<sup>†</sup> | 5,606 (2.8) | 21.0 (19.8, 22.2) | 5,311 (2.9) | 21.5 (20.3, 22.8) | 295 (1.8) | 11.5 (8.1, 15.9) |
| **Clinical severity\*** | Asymptomatic | 81,584 (40.9) | 19.9 (19.6, 20.2) | 77,057 (42.1) | 20.3 (19.9, 20.6) | 4,527 (28.0) | 14.1 (13.1, 15.3) |
|  | Mild/moderate | 7,602 (3.8) | 19.9 (18.9, 21.0) | 7,216 (3.9) | 20.1 (19.1, 21.2) | 386 (2.4) | 16.6 (12.9, 21.0) |
|  | Severe or critical | 32 (0.0) | 15.6 (5.9, 34.3) | 22 (0.0) | 13.6 (3.8, 36.4) | 10 (0.1) | 20.0 (4.0, 64.1) |

CI: confidence interval; <sup>†</sup> people of working age ( $\geq 18$ years) unemployed or not in the labor force (disabled); \*Clinical severity of first infection.

### 2.2 Vaccination after first Omicron variant infection

Figure 2 (Panel A) shows vaccination coverage in the analytic population over time. In April 2022, a series of emergency epidemic prevention measures, including mass screening with Nucleic Acid Amplification Tests (NAATs), city-wide lockdown, home quarantine of residents, were implemented in Shanghai, leading to the temporary suspension of vaccination in April and May 2022. By the end of the study period, January 2023, 69.5% (190,815/274,363) of the study participants had completed full vaccination, and 38.4% booster vaccination (**Figure S1)**. Vaccination coverages for the first, second, third, and fourth vaccine doses were 72.7%, 67.6%, 37.4% and 0.3%, respectively.

**Figure 2.**
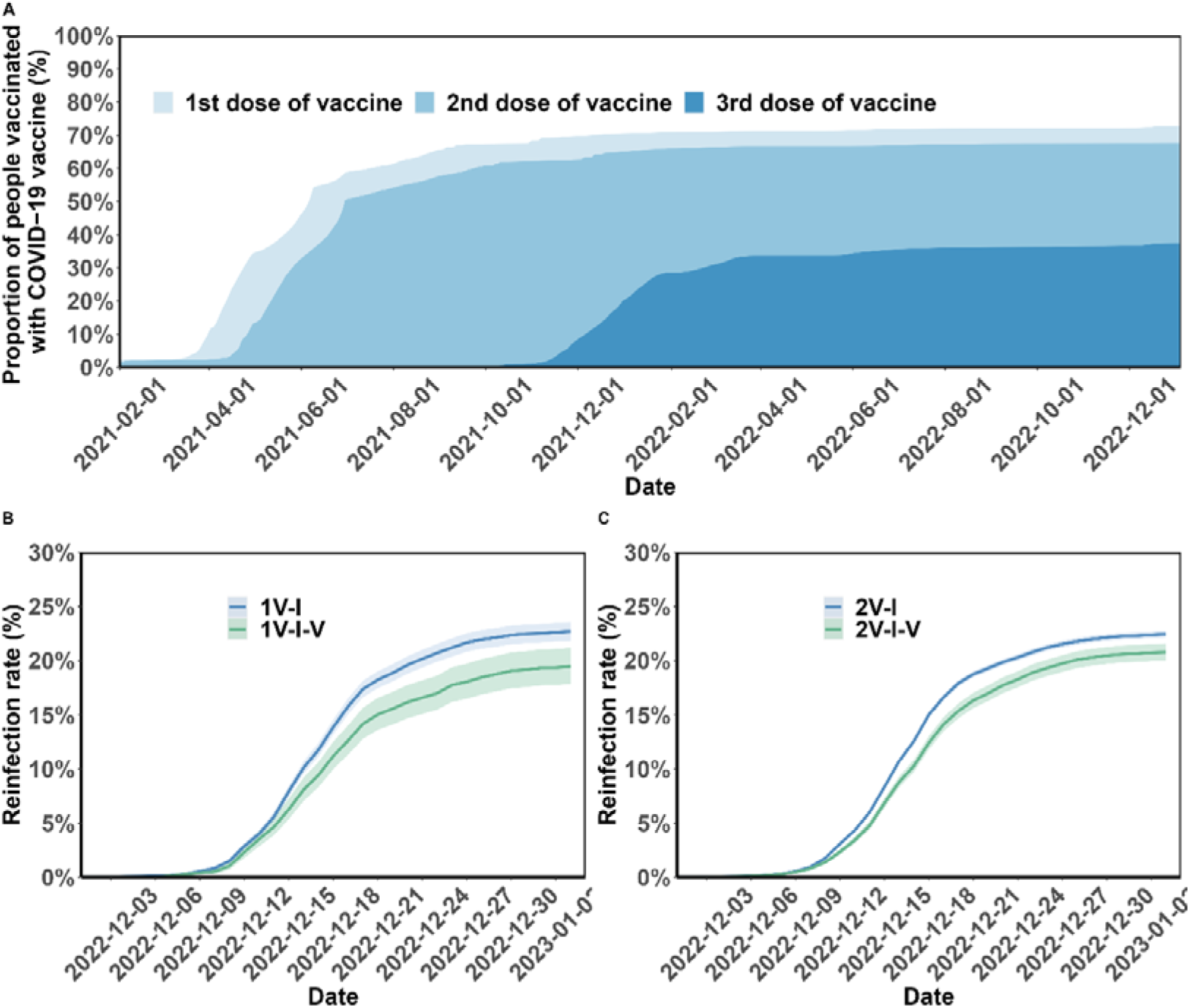
Vaccination coverage and cumulative incidence of SARS-CoV-2 reinfection in the study population. Panel A presents the percentages of the study population vaccinated over time. The cumulative incidence of SARS-CoV-2 reinfections is presented by the number of vaccination doses before (panels) and after (lines) first infection (panels B-C). Shaded regions: 95% CIs. 1V-I, 2V-I represented 1, and 2 vaccine doses before infection, respectively; 1V-I-V, 2V-I-V corresponds to 1 and 2 doses before infection, then post-infection vaccination, respectively. As mentioned in the *Results* section, 142 and 144 study participants who received one and two pre-infection vaccine doses received two post-infection vaccine doses. We do not show the corresponding plot for those individuals who received 3 pre-infection doses as their post-infection dose was after the start of the follow-up, in December.

As mentioned above, only participants who had received vaccination before the first infection were included in this analysis. After infection, 10,241 (5.1%), 5,096 (2.6%), and 810 (0.4%) individuals received another vaccine dose in August, and during the periods from September to November 2022 and from December 2022 to January 2023, respectively. Between their first infection, in April-May 2022, and January 2023, 17.4% (2,466/14,131) of the study participants who had one pre-infection dose received a second dose of COVID-19 vaccine, and 13.8% (12,886/93,087) of those who had received two pre-infection doses received a third vaccine dose. Only 1.0% (142/14,131) of the study participants who had one pre-infection dose received two post-infection vaccine doses, and 0.2% (144/93,087) of those who had received two pre-infection doses received two post-infection doses (**Table S2**). All individuals with three pre-infection vaccine doses who received a fourth dose (795/92,094, 0.9%) received the post-infection dose in December 2022; for 793 of these participants, the post-infection vaccination received in December was their second booster vaccination (for the other 2, their fourth dose was the first booster vaccination; see *Supplementary Appendix* for more information about vaccine policy in Shanghai).

### 2.3 SARS-CoV-2 reinfections

Among the study participants, 48,651/199,312 (24.4%) had SARS-CoV-2 reinfection. The median age of individuals with no evidence of reinfection was 41.7 years (IQR: 31.0, 55.7 years), and of individuals with confirmed SARS-CoV-2 reinfection, 45.8 years (IQR: 34.0, 55.9 years) (**Table S1**). The median time interval between the first infection and reinfection was 244.7 days (IQR: 237.4, 250.0), which implies that the risk of misclassifying a long infection as a reinfection was low.

Figure 2 (Panels B and C) shows the cumulative incidence of SARS-CoV-2 reinfection by vaccination status. Overall, individuals who were not vaccinated after their first infection were more often reinfected compared to those who received post-infection vaccination. The percentage of female individuals who became reinfected was lower than that of male individuals (22.4% *vs.* 26.1%; **Table 1**); and reinfection was more common in participants aged 40 to 59 years compared to those aged 20 to 39 (29.4% *vs.* 22.4%). Individuals originally from other provinces had a slightly higher risk of reinfection compared to individuals from Shanghai (20.9% vs. 18.9%). The risks for retired individuals, for those of working age who were not working, and for those working were 22.5, 21.0, and 20.8%, respectively.

### 2.4 Vaccine effectiveness

For individuals who had received at least one pre-infection vaccine dose, post-infection vaccination was protective against reinfection (adjusted hazard ratio [aHR] 0.82, 95% CI 0.79, 0.85). As shown in Figure 3, this protective effect was observed in subgroups defined by the number of pre-infection vaccine doses: aHR of 0.84 (95% CI, 0.76, 0.93) and 0.87 (95% CI, 0.83, 0.90) for one and two pre-infection doses respectively; and for patients with three vaccine doses prior to infection, the association was not statistically significant (aHR: 0.96 [0.74, 1.23]). When analyses are stratified by partial and full vaccination status before the first infection, post-infection vaccination was protective (aHR 0.76 [0.68, 0.84], and 0.93 [0.89, 0.97], respectively); and among individuals who had received booster vaccination before the first Omicron variant wave in Shanghai, the hazard ratio estimate was consistent with a more limited effect (aHR: 0.95 [0.75, 1.22]). Note that the overlapping categories between classification of pre-infection vaccination status and the number of pre-infection vaccinations (**Table S3)**. For comparison, results for individuals who had not been vaccinated before their first infection are shown in the *Supplementary Appendix* (supplementary section “Effect of post-infection vaccination in individuals with no history of vaccination before infection” and **Table S4**).

**Figure 3.**
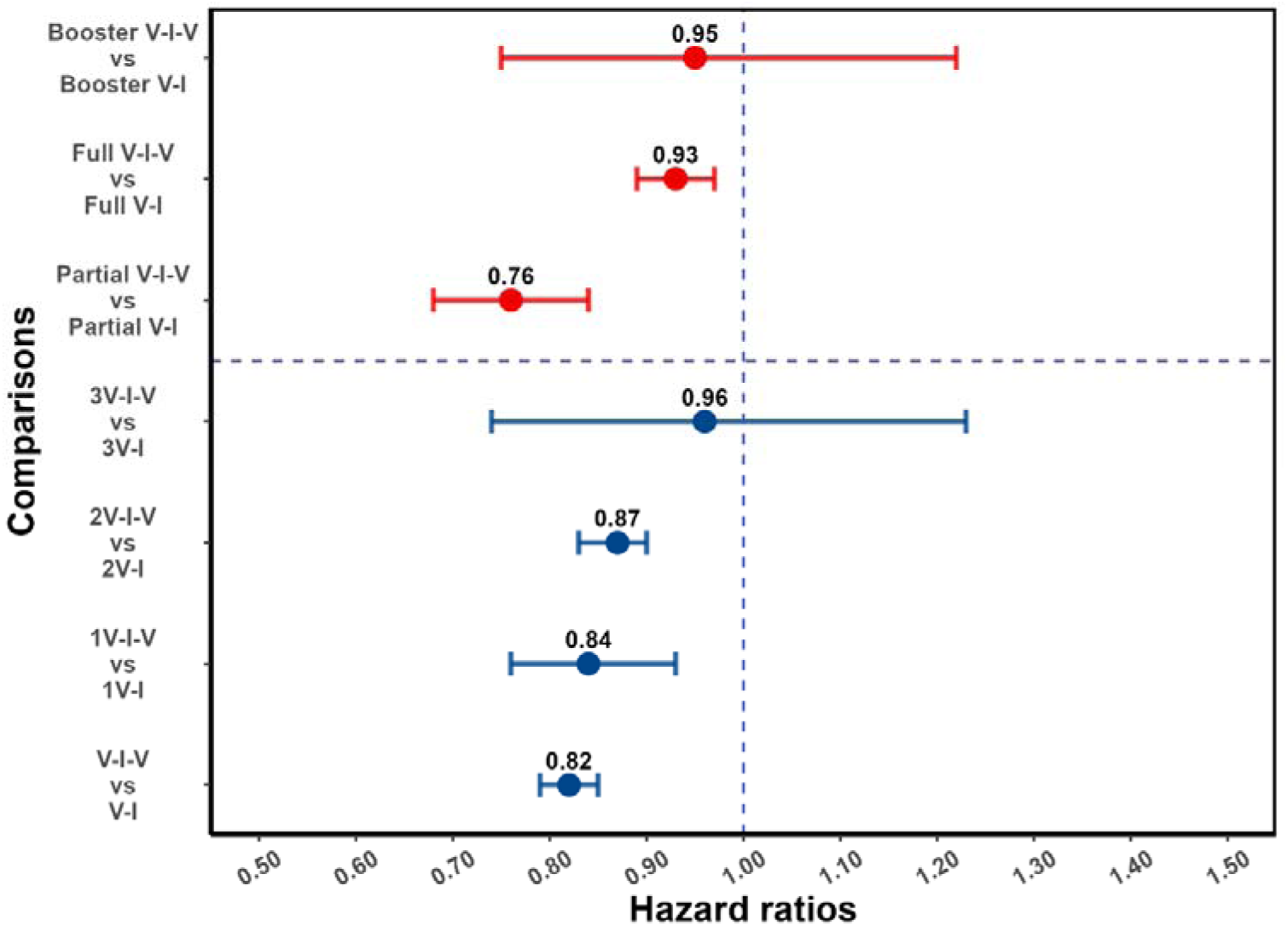
Effect of post-infection vaccination on SARS-CoV-2 reinfection stratified by pre-infection vaccination. Error bars (95% CIs) and circles represent aHR for SARS-CoV-2 reinfection estimated using Cox proportional hazards models. V-I-V, 1V-I-V, 2V-I-V, 3V-I-V corresponds to any pre-infection vaccination, 1, 2 and 3 vaccine doses before infection, then vaccination, respectively; they were compared to V-I, 1V-I, 2V-I, 3V-I, respectively. Partial V-I-V, Full V-I-V and Booster V-I-V represent partial vaccination, full vaccination and booster vaccination before infection, followed by post-infection vaccination, respectively. The number of doses received by individuals with partial versus full (and full with booster) vaccination depends on the type of SARS-CoV-2 vaccine received; in Table S3 we present a cross-classification of participants in the analytic population by these vaccination-related categorical variables.

In analyses stratified by demographic characteristics (Figure 4), post-infection vaccine doses were protective in both female (aHR 0.81 [0.76, 0.86]) and male individuals (aHR 0.83 [0.79, 0.87]). Post-infection vaccination was more protective for participants aged 60 years or older (aHR 0.73 [0.69, 0.78]) compared to other age groups. The estimated aHR for individuals who were asymptomatic during their first infection was 0.80 (0.74, 0.87), and for those who were symptomatic during the first infection was 1.01 (0.78, 1.29).

**Figure 4.**
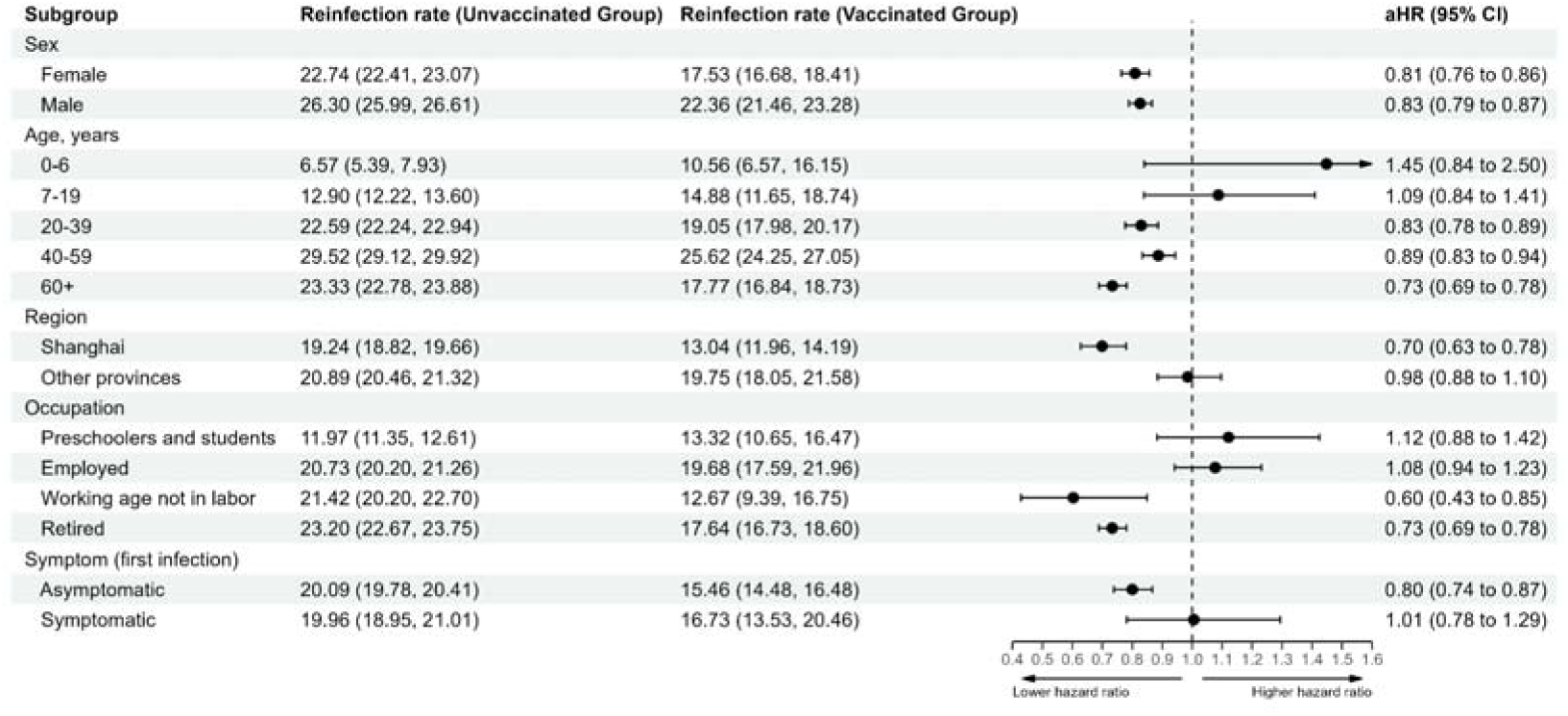
Vaccine-related protection against SARS-CoV-2 reinfection stratified by demographic characteristic. The vertical dotted line at 1.0 indicates no effect on protection.

As a secondary analysis, we estimated vaccine effects by calendar time of vaccination. Post-infection vaccine doses given within 30 and 90 days of the second Omicron variant wave were associated with lower hazard of reinfection (aHRs 0.51 [0.44, 0.58], 0.67 [0.61, 0.74], respectively). Note that in this secondary analysis, exposed and unexposed individuals were matched using propensity scores. We also performed a secondary analysis by vaccine type: for example, we compared patients who completed full vaccination before first infection and received a post-infection Ad5-nCOV vaccine dose, with those who completed full vaccination before first infection and did not receive post-infection vaccination; this approach was repeated for inactivated vaccines and recombinant protein vaccines (**Table S5**). In this analysis, the types of vaccines administered before infection often aligned between the post-infection vaccination group and the post-infection non-vaccination group. Post-infection Ad5-nCoV vaccine dose given to those had received full vaccination before the first infection was associated with lower hazard of reinfection (0.67, 95% CI: 0.56, 0.80); a protective effect was also observed for other vaccine types: inactivated vaccines (0.92, 95% CI: 0.87, 0.98) or recombinant protein vaccines (0.77, 95% CI: 0.64, 0.92). We performed an additional ad-hoc sensitivity analysis using a Cox model that was not adjusted for the severity of the first infection. As shown in **Figure S2**, our findings were not affected by the non-inclusion of disease severity in the survival analysis model.

## 3 Discussion

We aimed to estimate the added value of a vaccine dose given after SARS-CoV-2 infection in individuals who had received COVID-19 vaccination before the emergence of the Omicron variant. Although quantification of the protective effect of post-infection vaccination in individuals who have been both exposed to infection and previously vaccinated is a public health policy priority, as in many settings this group represents a large fraction of the population, studies on this question are often complicated by the variable timing of infection, vaccination and reinfection. Here, we leveraged the epidemiological history of COVID-19 in Shanghai municipality, where exposure to first and second SARS-CoV-2 infections occurred during well-defined periods of time, and large electronic databases allowed analyses of vaccination and infection history. Our study showed that post-infection vaccination reduced the risk of reinfection with the Omicron variant during a second, large surge of infections in the city. We also assessed the impact of the timing of vaccination relative to the second Omicron variant wave, and presented secondary and sensitivity analyses. This study will be used to guide COVID-19 vaccination policy in the municipality of Shanghai, and have implications for the rest of the country.

In Shanghai, COVID-19 vaccination started in February 2021 for individuals aged between 18 and 59 years, and over the following months, both older residents and children were included in the vaccination programme[9]. By the time of the first surge of Omicron variant infections, less than 70% of Shanghai residents aged 60 to 79 years had completed a primary vaccination series, and 40% had received a booster dose[8]. This was the context in which the population in this municipality was exposed to a first large epidemic of SARS-CoV-2 infections. The change in policy in December 2022 that was associated with a significant increase in the number of infections was our primary motivation to study the additional protection afforded by post-infection vaccination. Indeed, although epidemiological studies have been reported on the effect of vaccination against reinfection, primarily in Europe[10], North America[11] and Middle East[12], there is limited evidence from settings similar to that in Shanghai, where both the infection and vaccination histories, in terms of vaccine types, differ considerably from those in most Western countries. As individuals increasingly question the benefit of getting vaccinated after COVID-19 recovery, evidence generated locally is necessary.

Previous studies have shown that infection alone can result in immune responses that protect against Omicron variant reinfection. For example, a study in Qatar, with a test-negative design, found that an earlier Omicron variant infection provided protection against symptomatic BA.4 or BA.5 reinfection[13]. There are also different types of evidence that support post-infection vaccination. In Israel, post-infection vaccination protected against reinfection in individuals with no history of pre-infection vaccination[14], and prior SARS-CoV-2 infection was associated with lower risk for breakthrough infection among individuals receiving the BNT162b2 or mRNA-1273 vaccines in Qatar[15]. Further, a recent cohort study, performed in the US, of previously infected individuals who were unvaccinated at the time of first infection suggest that vaccination after recovery from COVID-19 decreased risk of reinfection by approximately half[16], which was consistent with a case-control study also conducted in the US[17]. Although our research question was different from those in these studies, data from Shanghai suggest that post-infection vaccination in individuals who had received at least one dose of SARS-CoV-2 vaccine before infection provides protection against reinfection. We note that whilst post-infection vaccination was protective for individuals who had received one or two vaccine doses before the first Omicron variant wave, for those individuals who had received three pre-infection vaccine doses, the evidence for a protective effect was limited. A possible explanation relates to the timing of post-infection vaccination: for all individuals with three pre-infection vaccine doses, the post-infection dose was given in December 2022 and, as a few weeks are required for immunologic boosting, the relatively short follow-up might have been insufficient to detect an effect. Similarly, evidence that post-infection vaccination was protective for individuals who had received booster vaccination before the spread of the first Omicron variant wave in Shanghai was limited; the majority (793/810) of these individuals had 3 doses before first Omicron variant infection.

In a secondary analysis that assessed the impact of the timing of the post-infection vaccination on the estimated effect, we observe that vaccination within 30 and 90 days before the second surge of Omicron variant provided different degrees of protection, consistent with waning of immunity even in individuals with multiple vaccine doses and history of infection[18]. As the duration of follow-up in our study was relatively short, we were not able to analyze immunity duration over longer periods of time after the start of the second Omicron variant wave. This observation has implications for COVID-19 vaccination programmes: for example, under perfectly functioning logistics, it is possible that timing of vaccination campaigns, including for individuals who have been both previously infected and vaccinated, might be optimal if launched immediately after increase in incidence (e.g. linked to a new variant), rather than immediately after the end of a wave of infections.

In our analyses stratified by demographic characteristics, the protective effect of post-infection vaccination was higher among patients who were 60 years of age or older than among younger individuals. As older individuals are more likely to suffer severe disease, including in settings similar to Shanghai[6], this finding suggests that vaccination of individuals in this age group, even after previous infection and vaccination, is beneficial. Similar to the findings of Ghorbani et al[19], we also found that the rate of reinfection in females was lower than in males; however, the protective effect of post-infection vaccination was similar in the two groups.

We also observed greater protection after an Ad5-nCoV post-infection vaccination in individuals who were fully vaccinated before their first SARS-CoV-2 infection compared to post-infection vaccination with other vaccine types. Note however that not all post-infection doses were of the same type of pre-infection vaccine doses. Another stratified analysis revealed that the protective effect of additional doses was different in individuals who had asymptomatic presentations of the first infection versus those who had symptomatic disease (aHR 0.80 [0.74, 0.87] and 1.01 [0.78, 1.29]). This could be related to different immune responses after infections with different severities. Indeed, reduced antibody response after the initial SARS-CoV-2 infection has been associated with incidence of reinfections. For example, in the study by Islamoglu and colleagues[20], it was observed that antibody responses against SARS-CoV-2 were protective against COVID-19 reinfection. Consistently, in the Republic of Korea, CD4^+^ T-cell responses tended to be greater in patients who had severe disease[21]. In agreement with these studies, that had different designs, our analysis suggests that individuals who had severe or critical COVID-19 had a lower risk of reinfection compared to those who were asymptomatic (see **Table 1**). Note that individuals who were asymptomatic and those with mild symptoms during the first infection had comparable rates of reinfection. However, individuals with unknown disease severity during the first infection had a higher reinfection rate compared to those with documented clinical severity.

Strengths of our study include: the well-defined timing of first infections and reinfections in Shanghai, linked to the two Omicron variant waves; detailed information on vaccination based on a citywide system; mass screening for SARS-CoV-2 infections during the first Omicron variant surge; free testing during the second Omicron variant wave. Further, to prevent immortal time bias, a common problem in epidemiological studies where treatment/exposure assignment is not aligned with the start of follow-up, we performed survival analysis that used a time varying exposure variable. The insights from our study are also likely generalizable to similar urban settings facing distinct waves of SARS-CoV-2 infections and that adopted the same vaccine types. However, our study also has limitations. Firstly, information on key variables, such as occupation, household registration, clinical severity during the first infection, was missing for a non-negligible fraction of the study population. Although in the primary analysis we performed adjustment for likely confounders, and in the secondary analysis we used propensity score matching, as in other observational studies, we cannot rule out the possibility of residual confounding, which in this context could be related to comorbidities that might affect both the decision to vaccinate after infection and risk of reinfection. Differences in healthcare-seeking behavior could also bias case ascertainment between post-infection vaccinated and unvaccinated individuals[22]. Although we restricted the study population to individuals who had received at least one pre-infection vaccination, which might have led to a higher degree of homogeneity in healthcare-seeking behaviour compared to that in the total population, it is possible that this bias might have affected our estimates. For example: individuals who were more health conscious might have been more likely to receive post-infection vaccination and also more likely to seek medical care or testing when reinfected, and this would have biased results toward the null; it is, however, also conceivable that these individuals were more likely to avoid contact with potentially infectious persons, which could have biased results in the opposite direction. Finally, data on the severity of infections during the second wave were not available, which prevented analyses of clinical outcomes other than infections (e.g. COVID-19-related hospitalization or death).

Although some previous studies [23–25] estimated similar or higher vaccine effectiveness against severe outcomes compared to outcomes that presumably include both milder and severe presentations, this pattern was not observed in all studies. Epidemiologists and public health officials who will use our results to define vaccination policy should thus take into account the fact that our analysis does not capture all benefits of post-infection vaccinations.

## Conclusions

Although SARS-CoV-2 vaccination, including booster doses, is recommended by the public health authorities in Shanghai to reduce the local disease burden caused by COVID-19, there is increasing unwillingness in the population to receive additional vaccine doses, and vaccine fatigue has been frequently reported. Our study provides evidence that there is additional value for individuals who have been vaccinated in receiving vaccine doses after infections. It also suggests that vaccination programmes need to be linked to efficient surveillance for new infections so that public health authorities can maximize impact of additional doses, including in this group of patients.

## 4 Materials and methods

### 4.1 Study setting and participants

During the first Omicron variant wave, the entire city of Shanghai entered a lockdown phase on April 1, 2022; and on June 1, 2022, the local government declared the end of the city-wide lockdown[6]. Residents (citizens, including immigrants from other provinces, and foreigners) in Shanghai, a provincial-level municipality in China with a population of more than 25 million people, underwent several rounds of SARS-CoV-2 RT-PCR testing between April 1 and May 31, 2022. This study included individuals diagnosed with their first SARS-CoV-2 infection between April 1 and May 31, 2022 in the Pudong District, which is a large and densely populated district of Shanghai spanning an area of 1,210 square kilometers with a permanent resident population of 5.57 million, served by more than 30 hospitals and 60 community health centers[26]; both individuals who were diagnosed by mass screening and those with symptoms who were seen by healthcare professionals were included. Information on infection history as well as data on demographic variables (sex at birth, and age) were provided by the Center for Disease Control and Prevention in Shanghai, China.

Additional information (e.g. on occupation, residence, clinical severity and symptoms of first infection) was available for patients with hospital records and those who were transferred to a hospital. The recorded clinical severity was categorized as asymptomatic, mild, moderate, severe, or critically ill[27, 28]. During the first Omicron variant wave in Shanghai, many Fangcang shelter hospitals were rapidly converted into facilities to treat COVID-19 patients and made important contributions in providing adequate healthcare to patients with mild to moderate symptoms, and preventing further viral transmission in the community[29]. The efficient referral and transfer mechanisms in the local communities meant that the majority of patients were admitted to Fangcang shelter hospitals one or two days after testing positive for SARS-CoV-2[30]. However, many patients were either admitted to hospital without complete clinical information or not transferred to other hospitals; for these study participants, information on clinical severity was often missing.

### 4.2 Study design and eligibility criteria

There was a second surge of Omicron variant cases from December 2022; and from January 2023, free nucleic acid testing services were no longer offered in Shanghai, and mandatory PCR testing on all personnel ceased. After this change, the cost of an individual test was 16 yuan (US$2.33)[31]. For this reason, the outcome variable in our analysis was based on reinfection data collected prior to January 2023. Reinfection-related death was defined as death within 30 days of a SARS-CoV-2 reinfection[10]; individuals who died from reasons unrelated to COVID-19 between the two Omicron variant waves were excluded. Individuals were also excluded if the date of first infection was missing. As the reasons why some individuals refused to receive vaccination varied and were often unknown[32], our analysis focused on individuals who had received at least one vaccine dose before the first SARS-CoV-2 infection.

### 4.3 Vaccination and reinfection data

The Shanghai Group Immunization System captures all vaccine administrations in the municipality and is updated daily. This system is linked to the National Immunization Program Information System, which also includes national identification-matched COVID-19 vaccinations received outside of Shanghai[8]. Vaccination status was categorized in accordance with national technical recommendations for COVID-19 vaccination[8]: unvaccinated, partially vaccinated, fully vaccinated and fully vaccinated with booster dose (here, also referred to as booster vaccination). Here, inactivated vaccines refer to Sinovac-CoronaVac, Sinopharm/BIBP COVID-19 vaccine, and Sinopharm/WIBP COVID-19 vaccine; Ad5-vectored vaccine refers to Cansino Ad5-nCoV-S COVID-19 vaccine; and recombinant protein vaccine refers to recombinant COVID-19 vaccine (CHO cell), Anhui Zhifei Longcom Biopharmaceutical Institute of Microbiology.

As in other epidemiological studies[19, 33], we defined reinfection as a positive SARS-CoV-2 RT-PCR or rapid antigen test at least 90 days after the first positive test. Phylogenetic analysis coupled with contact tracing data revealed community transmission of Omicron BA.5.2 sublineage in Shanghai[9]; the subvariant was estimated to have caused ∼90% of infections during the second Omicron variant wave.

### 4.4 Statistical analysis

Continuous variables are summarized as medians and interquartile ranges (IQR), and categorical variables, as counts, proportions or percentages. Cumulative incidence of reinfections was calculated and expressed as the number of reinfected individuals per 100 participants. Cox proportional hazards models with a time varying exposure variable corresponding to post-infection vaccination were used to estimate adjusted hazard ratios (aHR). In this analysis, time to reinfection was the outcome, and post-infection vaccination, the exposure of interest; models were adjusted for sex, age, residence, occupation and clinical severity of the first SARS-CoV-2 infection. As, due to social distancing measures, the number of SARS-CoV-2 infections diagnosed between May and November 2022 in Shanghai was low (N = 89 individuals), the start of the follow-up in the survival analysis was on December 1, and for each participant, the follow-up continued until January 3, 2023 or confirmed SARS-CoV-2 reinfection, whichever occurred earlier. We note that by using this approach, both the start of the follow-up and the eligibility (which required confirmed SARS-CoV-2 infection during the initial Omicron variant wave, and being at risk of infection when the second Omicron variant wave occurred in December) are temporally aligned. On the other hand, for some participants, the time of exposure (post-infection vaccination) occurred after time zero, which could potentially lead to immortal time bias[34]; for this reason, the exposure variable was as time-varying. We also assessed whether this effect was modified by demographic characteristics.

We conducted a secondary analysis to estimate the protection afforded by post-infection vaccination before the second Omicron variant wave (that is, only vaccine doses received before December 2022 were used in defining the exposure variable); this analysis was stratified by time intervals between post-infection vaccination and the second Omicron variant wave, and by post-infection vaccine type (for individuals who had the same vaccination status or the same number of doses before first infection). For this secondary analysis, propensity score matching was used to improve comparability between the exposed and unexposed groups; the propensity score was calculated using a logistic regression model with all available baseline characteristics, and a one-to-one matching was performed using the nearest neighbor matching method with a caliper width of 0.20[35]. We assessed the balance of covariates after matching using standardized mean differences (SMD), and considered a value of less than 0.1 to be indicative of adequate matching (**Table S6-8**). All statistical analyses were performed using R.4.1.1 software (Foundation for Statistical Computing, Vienna, Austria; https://www.r-project.org). We followed the Strengthening the reporting of observational studies in epidemiology (STROBE) recommendations, and the STROBE checklist is provided in the *Supplementary Appendix* (**Table S9**)

## Acknowledgments

We gratefully acknowledge all participants in this study.

## Additional information

### Funding

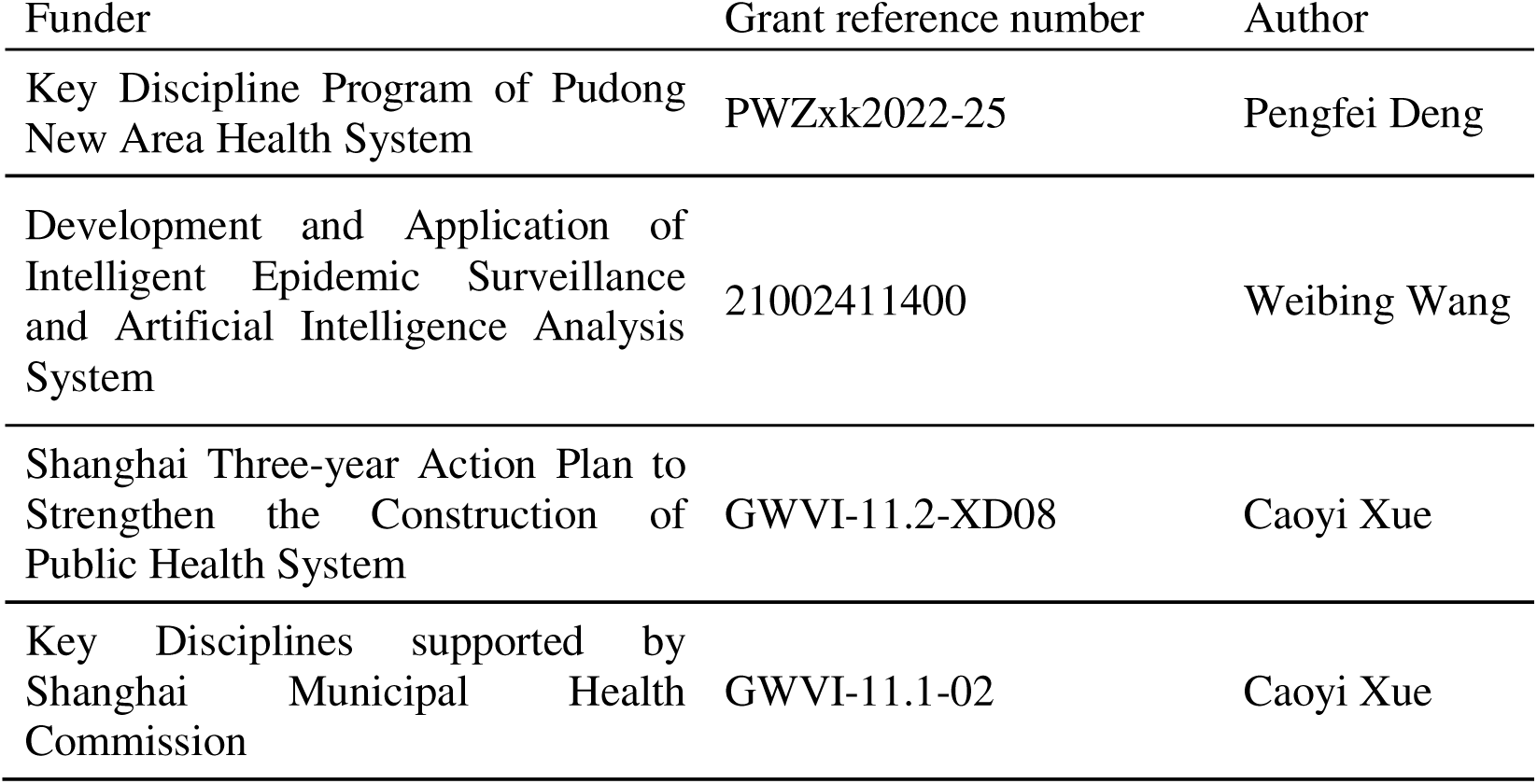
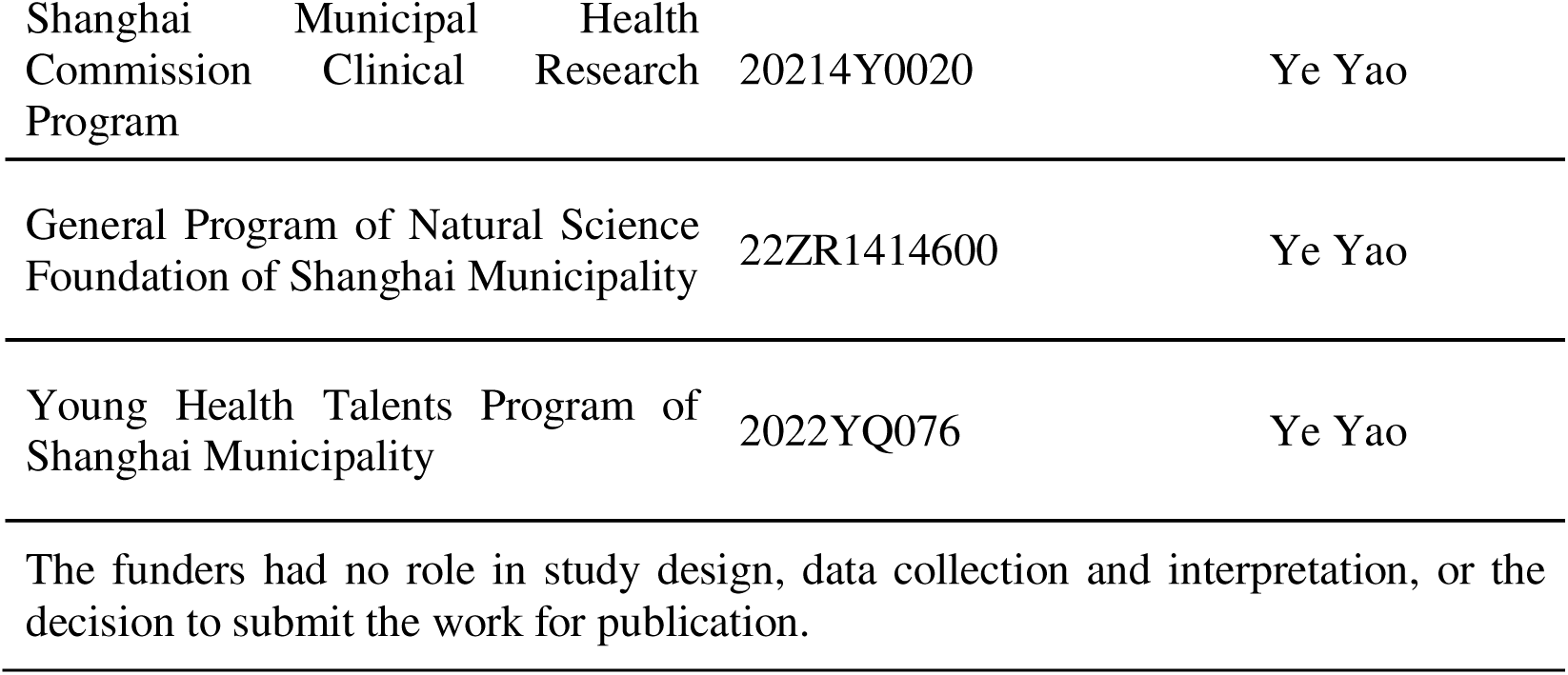

### Author contributions

BZ, BG, WW and YY designed the study. CX, PD and BZ extracted the data and constructed the database. BZ, JT and XL analyzed the data. BZ and WW drafted the manuscript—original draft. BG, XC, YY and WW conducted critical revisions of the manuscript—review and editing. All authors read and approved the final manuscript. **Ethics**

The study was approved by the Ethical Review Committee in the Shanghai Pudong New Area Center for Disease Control and Prevention (approval number: PDCDCLL-20220801-001).

### Consent for publication

Not applicable.

## Additional files

### Supplementary files

Table S1. Descriptive overview of the study population.

Table S2. Characteristics of vaccination distribution among study population.

Table S3. Cross-classification based on the number of vaccine doses received before the first infection and vaccination status.

Table S4. Effect of post-infection vaccination in individuals with no history of vaccination before infection.

Table S5. Effect of vaccination after first infection against SARS-CoV-2 reinfection stratified by sequence of vaccination dose and vaccination status before first infection. Table S6. Baseline demographic characteristics of unvaccinated and vaccinated groups after first infection stratified by dose received before first infection.

Table S7. Baseline demographic characteristics of unvaccinated and vaccinated groups after first infection stratified by intervals before the policy change.

Table S8. Baseline demographic characteristics of unvaccinated and vaccinated groups after first infection stratified by vaccination status prior infection.

Table S9. STROBE Statement.

Figure S1. Vaccination status coverage in the study population.

Figure S2. Effect of post-infection vaccination on SARS-CoV-2 reinfection stratified by pre-infection vaccination and not adjusted for the severity of the first infection.

## Data availability

The individual-level data used in this study are sensitive and cannot be publicly shared. Requests for data should be made to the Shanghai Pudong New Area Center for Disease Control and Prevention.

