## Supplementary Materials for "Protection afforded by post-infection SARS-CoV-2 vaccine doses: a cohort study in Shanghai" (from another version of this preprint: version 2, 2024-01-15)

**Summary**

Introduction of COVID-19 vaccination procedures

Table S1 to Table S7

Figure S1 to Figure S2

**Introduction of COVID-19 vaccination procedures**

Vaccination status was categorized into four levels in accordance with the national technical recommendations for COVID-19 vaccination: (1) unvaccinated—that is, no history of COVID-19 vaccination before first infection date; (2) partial vaccination—either one dose of inactivated vaccine, or two doses of recombinant protein vaccine (three doses are recommended for primary vaccination) before the first infection; (3) full primary vaccination—either two doses of inactivated vaccine, one dose of adenovirus vector vaccine, three doses of recombinant protein vaccine before first infection; or (4) booster vaccination—either two doses of Ad5-vectored vaccine; or two doses of inactivated vaccine and one booster dose of inactivated vaccine, or adenovirus vector vaccine; or three doses of recombinant protein and one booster dose of recombinant protein, or adenovirus vector vaccine before first infection date.

During the second Omicron wave in December 2022, Shanghai launched the implementation plan of the second dose of enhanced immunization of the novel coronavirus vaccine, encouraging second dose of enhanced immunization in high-risk groups: elderly people over 60 years old, people with serious underlying diseases and people with low immunity on the basis of the first dose of enhanced immunization.

**Table S1.** **Descriptive overview of the study population.** Whilst in Table 1 we present information by post-infection vaccination status, in this table data are shown based on reinfection during the second Omicron variant wave.

| **Characteristic** | | **N = 199,312 (%)** | **Not reinfection* (n = 150,661), n (%)** | **Reinfection^†^  (n = 48,651), n (%)** |
| --- | --- | --- | --- | --- |
| **Median age (IQR), years** | | 42.79 (31.77,55.73) | 41.66 (31.03,55.70) | 45.82 (33.97,55.85) |
| **Mean age (95% CI), years** | | 43.88 (43.8,43.95) | 43.37 (43.28,43.45) | 45.45 (45.32,45.58) |
| **Age group, years** | **0–6** | 1736 (0.9) | 1615 (1.1) | 121 (0.2) |
|  | **7–19** | 10762 (5.4) | 9363 (6.2) | 1399 (2.9) |
|  | **20–39** | 75955 (38.1) | 58911 (39.1) | 17044 (35.0) |
|  | **40–59** | 74680 (37.5) | 52726 (35.0) | 21954 (45.1) |
|  | **≥ 60** | 35903 (18.0) | 27796 (18.4) | 8107 (16.7) |
| **Sex** | **Male** | 112672 (56.5) | 83263 (55.3) | 29409 (60.4) |
|  | **Female** | 85804 (43.1) | 66581 (44.2) | 19223 (39.5) |
| **Vaccination status** | **Partial vaccination** | 8497 (4.3) | 6968 (4.6) | 1529 (3.1) |
|  | **Full vaccination** | 85546 (42.9) | 66486 (44.2) | 19060 (39.2) |
|  | **Booster vaccination** | 105269 (52.8) | 77207 (51.2) | 28062 (57.7) |
| **Clinical severity during the first infection** | **Asymptomatic** | 81584 (40.9) | 65333 (43.4) | 16251 (33.4) |
|  | **Mild and moderate** | 7602 (3.8) | 6087 (4.0) | 1515 (3.1) |
|  | **Severe or critical** | 32 (0.0) | 27 (0.0) | 5 (0.0) |

IQR: interquartile range. Data are presented as means with 95% confidence intervals (95% CIs) or as proportions (%). Note that for the variables on age, sex, and clinical severity, data are missing for fractions of the study population, especially the clinical severity during the first infection.

**Table S2. Baseline demographic characteristics of unvaccinated and vaccinated groups after first infection stratified by dose received before first infection.** This comparison only refers to the secondary analyses where exposed and unexposed individuals were matched based on propensity scores.

| **Characteristics** | | **Received 1 vaccine dose prior first infection** | | | **Received 2 vaccine doses prior first infection** | | |
| --- | --- | --- | --- | --- | --- | --- | --- |
|  |  | **Unvaccinated group (N = 2,247)** | **Vaccinated group (N = 2,247)** | **SMD** | **Unvaccinated group (N = 12,674)** | **Vaccinated group (N = 12,674)** | **SMD** |
| **Sex** | **Male** | 1309 (58.3) | 1309 (58.3) | <0.001 | 6784 (53.5) | 6784 (53.5) | <0.001 |
|  | **Female** | 924 (41.1) | 924 (41.1) | | 5841 (46.1) | 5841 (46.1) | |
|  | **Unknown** | 14 (0.6) | 14 (0.6) |  | 49 (0.4) | 49 (0.4) |  |
| **Age, years** | **0-6** | 78 (3.5) | 78 (3.5) | <0.001 | 89 (0.7) | 89 (0.7) | <0.001 |
|  | **7-19** | 155 (6.9) | 155 (6.9) | | 258 (2.0) | 258 (2.0) | |
|  | **20-39** | 819 (36.4) | 819 (36.4) | | 3828 (30.2) | 3828 (30.2) | |
|  | **40-59** | 479 (21.3) | 479 (21.3) | | 3323 (26.2) | 3323 (26.2) | |
|  | **60+** | 709 (31.6) | 709 (31.6) | | 5142 (40.6) | 5142 (40.6) | |
|  | **Unknown** | 7 (0.3) | 7 (0.3) |  | 34 (0.3) | 34 (0.3) |  |
| **Regions** | **Shanghai** | 370 (16.5) | 370 (16.5) | <0.001 | 2369 (18.7) | 2369 (18.7) | <0.001 |
|  | **Other provinces** | 296 (13.2) | 296 (13.2) | | 1481 (11.7) | 1481 (11.7) | |
|  | **Unknown** | 1581 (70.4) | 1581 (70.4) | | 8824 (69.6) | 8824 (69.6) | |
| **Occupations** | **Preschoolers and students** | 228 (10.1) | 228 (10.1) | <0.001 | 326 (2.6) | 326 (2.6) | <0.001 |
|  | **Employed** | 161 (7.2) | 161 (7.2) | | 865 (6.8) | 865 (6.8) | |
|  | **Working age not in labor** | 31 (1.4) | 31 (1.4) |  | 245 (1.9) | 245 (1.9) | |
|  | **Retired** | 716 (31.9) | 716 (31.9) | | 5205 (41.1) | 5205 (41.1) | |
|  | **Unknown** | 1111 (49.4) | 1111 (49.4) | | 6033 (47.6) | 6033 (47.6) | |
| **Clinical severity** | **Asymptomatic** | 612 (27.2) | 612 (27.2) | 0.043 | 3546 (28.0) | 3546 (28.0) | 0.016 |
|  | **Mild and moderate** | 54 (2.4) | 52 (2.3) |  | 301 (2.4) | 297 (2.3) | |
|  | **Severe or critical** | 0 (0.0) | 2 (0.1) |  | 3 (0.0) | 7 (0.1) |  |
|  | **Unknown** | 1581 (70.4) | 1581 (70.4) | | 8824 (69.6) | 8824 (69.6) | |

SMD standardized mean difference.

**Table S3. Baseline demographic characteristics of unvaccinated and vaccinated groups after first infection stratified by intervals before the policy change.** This comparison only refers to the secondary analyses where exposed and unexposed individuals were matched based on propensity scores.

| **Characteristics** | | **Within 30 days*** | | | **Within 90 days*** | | | | |
| --- | --- | --- | --- | --- | --- | --- | --- | --- | --- |
|  |  | **Unvaccinated group (N = 3,137)** | **Vaccinated group (N =3,137)** | **SMD** | **Unvaccinated group (N = 4,161)** | | **Vaccinated group (N = 4,161)** | | **SMD** |
| **Sex** | **Male** | 1585 (50.5) | 1585 (50.5) | <0.001 | 2275 (54.7) | 2275 (54.7) | | <0.001 | |
|  | **Female** | 1541 (49.1) | 1541 (49.1) |  | 1865 (44.8) | 1865 (44.8) | |  | |
|  | **Unknown** | 11 (0.4) | 11 (0.4) |  | 21 (0.5) | 21 (0.5) | |  | |
| **Age, years** | **0-6** | 69 (2.2) | 69 (2.2) | <0.001 | 77 (1.9) | 77 (1.9) | | <0.001 | |
|  | **7-19** | 56 (1.8) | 56 (1.8) |  | 107 (2.6) | 107 (2.6) | |  | |
|  | **20-39** | 870 (27.7) | 870 (27.7) |  | 1472 (35.4) | 1472 (35.4) | |  | |
|  | **40-59** | 645 (20.6) | 645 (20.6) |  | 1003 (24.1) | 1003 (24.1) | |  | |
|  | **60+** | 1495 (47.7) | 1495 (47.7) |  | 1494 (35.9) | 1494 (35.9) | |  | |
|  | **Unknown** | 2 (0.1) | 2 (0.1) |  | 8 (0.2) | 8 (0.2) | |  | |
| **Regions** | **Shanghai** | 1015 (32.4) | 1015 (32.4) | <0.001 | 1060 (25.5) | 1060 (25.5) | | <0.001 | |
|  | **Other provinces** | 341 (10.9) | 341 (10.9) |  | 646 (15.5) | 646 (15.5) | |  | |
|  | **Unknown** | 1781 (56.8) | 1781 (56.8) |  | 2455 (59.0) | 2455 (59.0) | |  | |
| **Occupations** | **Preschoolers and students** | 124 (4.0) | 124 (4.0) | <0.001 | 180 (4.3) | 180 (4.3) | | <0.001 | |
|  | **Employed** | 181 (5.8) | 181 (5.8) |  | 344 (8.3) | 344 (8.3) | |  | |
|  | **Working age not in labor** | 54 (1.7) | 54 (1.7) |  | 81 (1.9) | 81 (1.9) | |  | |
|  | **Retired** | 1526 (48.6) | 1526 (48.6) |  | 1528 (36.7) | 1528 (36.7) | |  | |
|  | **Unknown** | 1252 (39.9) | 1252 (39.9) |  | 2028 (48.7) | 2028 (48.7) | |  | |
| **Clinical severity** | **Asymptomatic** | 1273 (40.6) | 1273 (40.6) | 0.01 | 1600 (38.5) | 1600 (38.5) | | <0.001 | |
|  | **Mild and moderate** | 79 (2.5) | 80 (2.6) |  | 102 (2.5) | 102 (2.5) | |  | |
|  | **Severe or critical** | 4 (0.1) | 3 (0.1) |  | 4 (0.1) | 4 (0.1) | |  | |
|  | **Unknown** | 1781 (56.8) | 1781 (56.8) |  | 2455 (59.0) | 2455 (59.0) | |  | |

SMD standardized mean difference. *before second Omicron variant wave

**Table S4. Baseline demographic characteristics of unvaccinated and vaccinated groups after first infection stratified by vaccination status prior infection.** This comparison only refers to the secondary analyses where exposed and unexposed individuals were matched based on propensity scores.

| **Characteristics** | | **Partial vaccination prior to infection** | | | | **Full vaccination prior to infection** | | |
| --- | --- | --- | --- | --- | --- | --- | --- | --- |
|  |  | **Unvaccinated group (N = 2,451)** | **Vaccinated group (N = 2,451)** | **SMD** | **Unvaccinated group (N = 11,034)** | | **Vaccinated group (N = 11,034)** | **SMD** |
| **Sex** | **Male** | 1286 (52.5) | 1286 (52.5) | <0.001 | 6133 (55.6) | | 6133 (55.6) | <0.001 |
|  | **Female** | 1148 (46.8) | 1148 (46.8) |  | 4859 (44.0) | | 4859 (44.0) |  |
|  | **Unknown** | 17 (0.7) | 17 (0.7) |  | 42 (0.4) | | 42 (0.4) |  |
| **Age, years** | **0-6** | 163 (6.7) | 163 (6.7) | <0.001 | 181 (1.6) | | 181 (1.6) | <0.001 |
|  | **7-19** | 232 (9.5) | 232 (9.5) |  | 3722 (33.7) | | 3722 (33.7) |  |
|  | **20-39** | 905 (36.9) | 905 (36.9) |  | 3186 (28.9) | | 3186 (28.9) |  |
|  | **40-59** | 559 (22.8) | 559 (22.8) |  | 3914 (35.5) | | 3914 (35.5) |  |
|  | **60+** | 585 (23.9) | 585 (23.9) |  | 31 (0.3) | | 31 (0.3) |  |
|  | **Unknown** | 7 (0.3) | 7 (0.3) |  | 1674 (15.2) | | 1674 (15.2) | <0.001 |
| **Regions** | **Shanghai** | 460 (18.8) | 460 (18.8) | <0.001 | 1428 (12.9) | | 1428 (12.9) |  |
|  | **Other provinces** | 330 (13.5) | 330 (13.5) |  | 7932 (71.9) | | 7932 (71.9) |  |
|  | **Unknown** | 1661 (67.8) | 1661 (67.8) |  | 168 (1.5) | | 168 (1.5) | <0.001 |
| **Occupations** | **Preschoolers and students** | 382 (15.6) | 382 (15.6) | <0.001 | 834 (7.6) | | 834 (7.6) |  |
|  | **Employed** | 184 (7.5) | 184 (7.5) |  | 186 (1.7) | | 186 (1.7) |  |
|  | **Working age not in labor** | 62 (2.5) | 62 (2.5) |  | 3959 (35.9) | | 3959 (35.9) |  |
|  | **Retired** | 593 (24.2) | 593 (24.2) |  | 5887 (53.4) | | 5887 (53.4) |  |
|  | **Unknown** | 1230 (50.2) | 1230 (50.2) |  | 2827 (25.6) | | 2827 (25.6) | <0.001 |
| **Clinical severity** | **Asymptomatic** | 726 (29.6) | 726 (29.6) | 0.039 | 272 (2.5) | | 272 (2.5) |  |
|  | **Mild and moderate** | 63 (2.6) | 60 (2.4) |  | 3 (0.0) | | 3 (0.0) |  |
|  | **Severe or critical** | 1 (0.0) | 4 (0.2) |  | 7932 (71.9) | | 7932 (71.9) |  |
|  | **Unknown** | 1661 (67.8) | 1661 (67.8) |  | 7943 (71.9) | | 7943 (71.9) |  |

SMD standardized mean difference.

**Table S5. Characteristics of vaccination distribution among study population.**

| **Number of vaccine doses before infection** | **Participants in the analysis** | **Participants received post-infection dose** | **1 dose after infection** | | **2 doses after infection** | | |
| --- | --- | --- | --- | --- | --- | --- | --- |
|  | **Total** | **Total** | **Total** | **Before Dec.** | **Total** | **Dose 1 before Dec.** | **Dose 2 before Dec.** |
| 1 | 14131 | 2466 | 2324 | 2324 | 142 | 142 | 13 |
| 2 | 93087 | 12886 | 12742 | 12727 | 144 | 144 | 32 |
| 3 | 92094 | 795 | 795 | 0 | 0 | 0 | 0 |

**Table S6. Effect of vaccination after first infection against SARS-CoV-2 reinfection stratified by sequence of vaccination dose and vaccination status before first infection.**

| **Characteristics**^#^ | | **Unvaccinated group** | | **Vaccinated group** | | **Number with same type of vaccine before infection, N (%)** | **aHR (95% CI)** |
| --- | --- | --- | --- | --- | --- | --- | --- |
|  |  | **N (%)** | **Reinfection rate, % (95% CI)** | **N (%)** | **Reinfection rate, % (95% CI)** |  |  |
| **1 dose of vaccine before first infection** | **Overall** | 2247 (100) | 23.63 (21.69, 25.71) | 2247 (100) | 19.94 (18.16, 21.85) | 1284 (57.14) | 0.81 (0.72, 0.92) |
|  | **Ad5-nCoV** | 423 (18.83) | 27.90 (23.20, 33.28) | 423 (18.83) | 26.00 (21.48, 31.21) | 213 (50.35) | 0.91 (0.70, 1.18) |
|  | **Inactivated vaccine** | 1679 (74.72) | 22.75 (20.56, 25.12) | 1679 (74.72) | 18.23 (16.27, 20.35) | 1064 (63.37) | 0.77 (0.66, 0.89) |
|  | **Recombinant protein vaccine** | 12 (0.53) | 33.33 (11.14, 79.25) | 12 (0.53) | 41.67 (15.80, 91.33) | 7 (58.33) | 1.28 (0.34, 4.80) |
| **2 doses of vaccine before first infection** | **Overall** | 12674 (100) | 24.57 (23.72, 25.44) | 12674 (100) | 20.81 (20.03, 21.62) | 11979 (94.52) | 0.83 (0.79, 0.87) |
|  | **Ad5-nCoV** | 787 (6.21) | 24.27 (21.01, 27.90) | 787 (6.21) | 11.61 (9.40, 14.20) | 512 (65.06) | 0.45 (0.35, 0.58) |
|  | **Inactivated vaccine** | 10747 (84.80) | 24.60 (23.68, 25.55) | 10747 (84.80) | 21.39 (20.53, 22.27) | 10465 (97.38) | 0.85 (0.80, 0.90) |
|  | **Recombinant protein vaccine** | 1057 (8.34) | 25.26 (22.37, 28.43) | 1057 (8.34) | 20.91 (18.29, 23.80) | 1002 (94.80) | 0.79 (0.66, 0.95) |
| **Partial vaccination before first infection**^†^ | **Overall** | 2451 (100) | 20.52 (18.79, 22.38) | 2451 (100) | 16.40 (14.86, 18.06) | 2349 (95.84) | 0.78 (0.69, 0.89) |
|  | **Inactivated vaccine** | 2343 (95.59) | 20.44 (18.67, 22.34) | 2343 (95.59) | 16.30 (14.73, 18.00) | 2304 (98.34) | 0.78 (0.68, 0.90) |
|  | **Recombinant protein vaccine** | 47 (1.92) | 19.15 (9.46, 34.95) | 47 (1.92) | 17.02 (8.03, 32.12) | 45 (95.74) | 0.92 (0.35, 2.38) |
| **Full vaccination before first infection** | **Overall** | 11034 (100) | 25.33 (24.40, 26.28) | 11034 (100) | 22.72 (21.84, 23.62) | 9542 (86.48) | 0.88 (0.83, 0.93) |
|  | **Ad5-nCoV** | 1209 (10.96) | 24.32 (21.66, 27.22) | 1209 (10.96) | 17.12 (14.91, 19.58) | 805 (66.58) | 0.67 (0.56, 0.80) |
|  | **Inactivated vaccine** | 8787 (79.64) | 25.37 (24.33, 26.44) | 8787 (79.64) | 23.69 (22.69, 24.73) | 7944 (90.41) | 0.92 (0.87, 0.98) |
|  | **Recombinant protein vaccine** | 1037 (9.40) | 26.13 (23.16, 29.39) | 1037 (9.40) | 20.93 (18.28, 23.85) | 793 (76.47) | 0.77 (0.64, 0.92) |

Data are presented as average (95%CI), or n (n/N%), where N is the total number of patients. ^#^ individuals received two doses of inactivated vaccine, recombinant protein vaccine or Ad5-nCoV were excluded. †only few post-infection Ad5-nCoV vaccine dose given to those had partial vaccination before the first infection

**Table S7. STROBE Statement**—Checklist of items that should be included in reports of ***cohort studies***

|  | Item No | Recommendation | Page/Figure |
| --- | --- | --- | --- |
| Title and abstract | 1 | (a) Indicate the study’s design with a commonly used term in the title or the abstract | 1 |
|  |  | (b) Provide in the abstract an informative and balanced summary of what was done and what was found | 1 |
| Introduction | | |  |
| Background/rationale | 2 | Explain the scientific background and rationale for the investigation being reported | 2 |
| Objectives | 3 | State specific objectives, including any prespecified hypotheses | 2 |
| Methods | | |  |
| Study design | 4 | Present key elements of study design early in the paper | 3 |
| Setting | 5 | Describe the setting, locations, and relevant dates, including periods of recruitment, exposure, follow-up, and data collection | 2-3 |
| Participants | 6 | (a) Give the eligibility criteria, and the sources and methods of selection of participants. Describe methods of follow-up | 2-3 |
|  |  | (b) For matched studies, give matching criteria and number of exposed and unexposed | 4-5 |
| Variables | 7 | Clearly define all outcomes, exposures, predictors, potential confounders, and effect modifiers. Give diagnostic criteria, if applicable | 4 |
| Data sources/ measurement | 8* | For each variable of interest, give sources of data and details of methods of assessment (measurement). Describe comparability of assessment methods if there is more than one group | 4 |
| Bias | 9 | Describe any efforts to address potential sources of bias | 4-5 |
| Study size | 10 | Explain how the study size was arrived at |  |
| Quantitative variables | 11 | Explain how quantitative variables were handled in the analyses. If applicable, describe which groupings were chosen and why | 4-5 |
| Statistical methods | 12 | (a) Describe all statistical methods, including those used to control for confounding | 4-5 |
|  |  | (b) Describe any methods used to examine subgroups and interactions | 4-5 |
|  |  | (c) Explain how missing data were addressed | 4-5 |
|  |  | (d) If applicable, explain how loss to follow-up was addressed | - |
|  |  | (e) Describe any sensitivity analyses | 4-5 |
| Results | | |  |
| Participants | 13* | (a) Report numbers of individuals at each stage of study—eg numbers potentially eligible, examined for eligibility, confirmed eligible, included in the study, completing follow-up, and analyzed | 2-3 |
|  |  | (b) Give reasons for non-participation at each stage | 3 |
|  |  | (c) Consider use of a flow diagram | Figure 1 |
| Descriptive data | 14* | (a) Give characteristics of study participants (eg demographic, clinical, social) and information on exposures and potential confounders | 5-6 |
|  |  | (b) Indicate number of participants with missing data for each variable of interest | 5-6 |
|  |  | (c) Summarize follow-up time (eg, average and total amount) | 4-5 |
| Outcome data | 15* | Report numbers of outcome events or summary measures over time | Table1 and Table S1 |
| \Main results | 16 | (a) Give unadjusted estimates and, if applicable, confounder-adjusted estimates and their precision (eg, 95% confidence interval). Make clear which confounders were adjusted for and why they were included | 7-8 |
|  |  | (b) Report category boundaries when continuous variables were categorized | Figure 2-4 |
|  |  | (c) If relevant, consider translating estimates of relative risk into absolute risk for a meaningful time period | 5-8 |
| Other analyses | 17 | Report other analyses done—eg analyses of subgroups and interactions, and sensitivity analyses | 8 |
| Discussion | | |  |
| Key results | 18 | Summarize key results with reference to study objectives | 8-9 |
| Limitations | 19 | Discuss limitations of the study, taking into account sources of potential bias or imprecision. Discuss both direction and magnitude of any potential bias | 11 |
| Interpretation | 20 | Give a cautious overall interpretation of results considering objectives, limitations, multiplicity of analyses, results from similar studies, and other relevant evidence | 9-11 |
| Generalisability | 21 | Discuss the generalisability (external validity) of the study results | 12 |
| Other information | | |  |
| Funding | 22 | Give the source of funding and the role of the funders for the present study and, if applicable, for the original study on which the present article is based | 12 |

*Give information separately for exposed and unexposed groups.

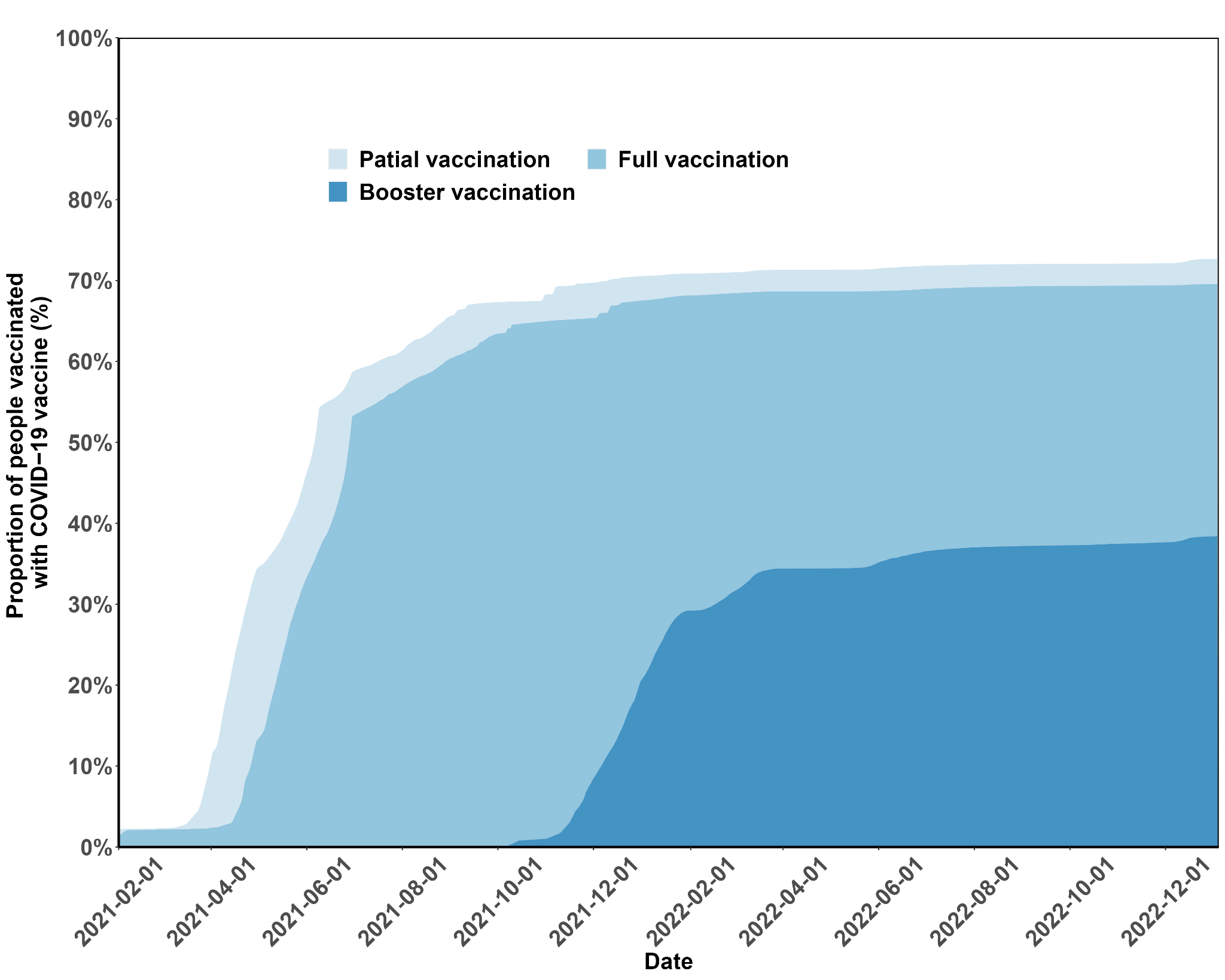

**Figure S1. Vaccination status coverage in the study population.** The figure presents the percentages of the study population vaccinated over time.

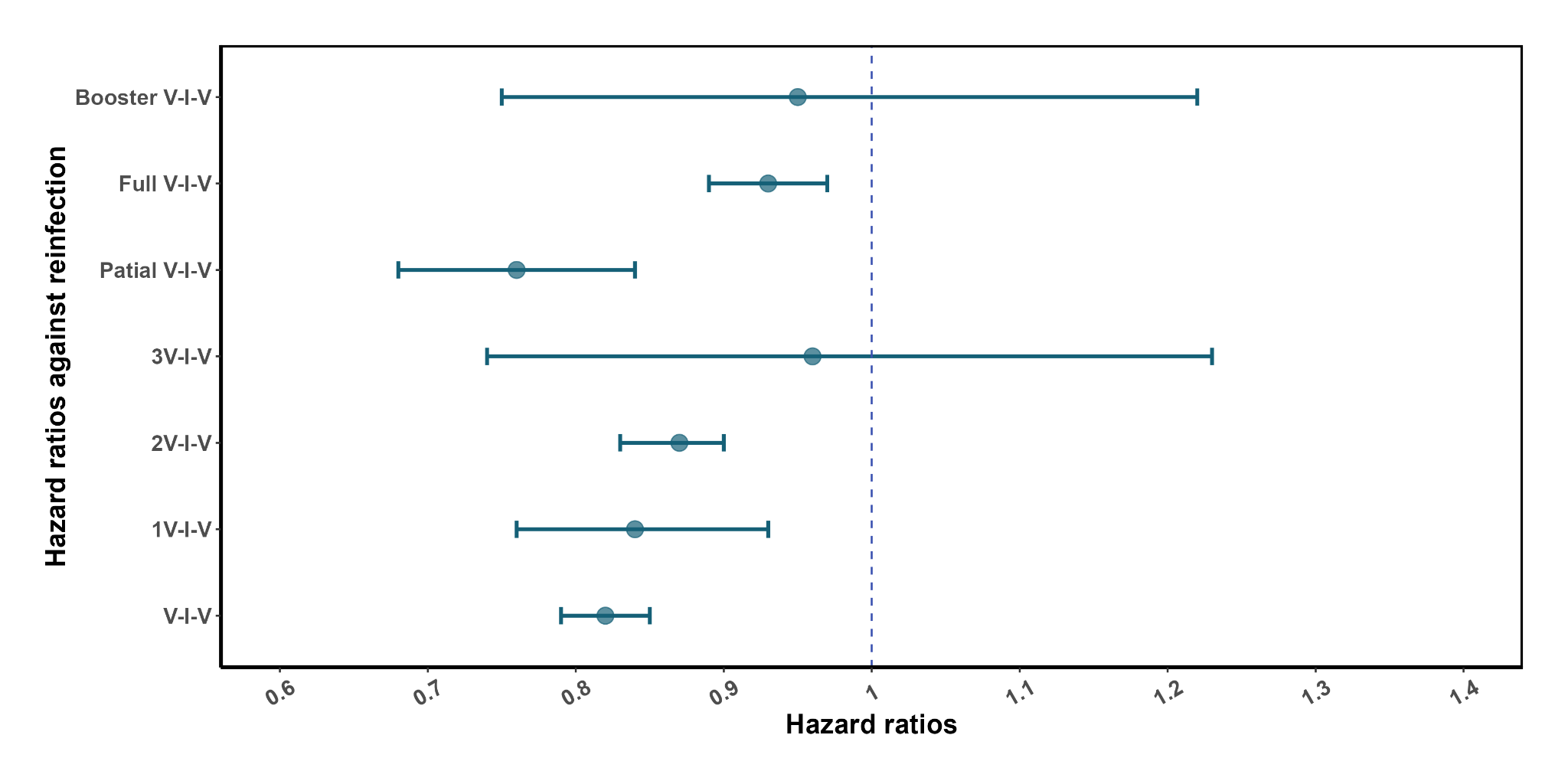

**Figure S2. Effect of vaccine dose on SARS-CoV-2 reinfection stratified by pre-infection vaccination and not adjusted for the severity of the first infection.** Error bars (95% CIs) and dots represent aHR for SARS-CoV-2 reinfection estimated using Cox proportional hazards models. 1V-I-V, 2V-I-V, 3V-I-V corresponds to 1, 2 and 3 vaccine doses before infection, then vaccination, respectively; they were compared to 1V-I, 2V-I, 3V-I, respectively. V-I-V, Partial V-I-V, Full V-I-V and Booster V-I-V represent any pre-infection vaccination, partial vaccination, full vaccination and booster vaccination before infection, followed by post-infection vaccination, respectively.
